# Race/ethnicity and socioeconomic status affect the assessment of lipoprotein(a) levels in clinical practice

**DOI:** 10.1101/2024.05.14.24307362

**Authors:** Marianna Pavlyha, Yihao Li, Sarah Crook, Brett R. Anderson, Gissette Reyes-Soffer

**Affiliations:** Department of Medicine, Columbia University Vagelos College of Physicians and Surgeons, New York, NY; Department of Neurology, Columbia University Vagelos College of Physicians and Surgeons New York, NY; Department of Pediatrics Cardiology, Columbia University Vagelos College of Physicians and Surgeons, New York, NY; Mindich Child Health and Development Institute, Icahn School of Medicine at Mount Sinai, New York, NY; Department of Population Health Sciences and Policy, Icahn School of Medicine at Mount Sinai, New York, NY; Department of Pediatrics, Icahn School of Medicine at Mount Sinai, New York, NY

**Keywords:** Lipoprotein(a), lipoproteins, ASCVD, cardiovascular disease, socioeconomic score, racial disparities

## Abstract

**Background and Objective:** High Lp(a) levels are a risk factor for ASCVD, however Lp(a) ordering in clinical practice is low. This study examines how race/ethnicity and socioeconomic status influence Lp(a) ordering.

**Methods:** This is a single center, retrospective study (2/1/2020-6/30/2023) using electronic medical records of adults with at least one ICD-10 diagnosis of ASCVD or resistant hyperlipidemia (LDL-C >160 mg/dL on statin therapy). We evaluated Lp(a) level differences among racial/ethnic groups and sexes. We also assessed associations between diagnosis type, diagnosis number, age at diagnosis, race, socioeconomic score (based on zip codes), public health coverage and presence of Lp(a) orders.

**Results:** 4% of our cohort (N=56,833) had an Lp(a) order (17.3% Hispanic, 8.7% non-Hispanic Black, 47.5% non-Hispanic White and, 27% Asian/others). Non-Hispanic Black and Hispanic patients had lower rates of Lp(a) orders (0.17%, 0.28%, respectively) when compared to non-Hispanic White patients (2.35%), p<0.001, however, their median Lp(a) levels were higher. Individuals belonging to deprived socioeconomic groups or on Medicaid, were less likely to have an Lp(a) order (RR=0.39, p<0.001 and RR=0.40, p<0.001 respectively). Certain diagnoses (carotid stenosis, family history of ASCVD and FH) and multiple diagnoses (>2) resulted in more Lp(a) orders compared to those with only one diagnosis (p<0.001).

**Conclusions:** Lp(a) ordering is low in patients with ASCVD. Non-Hispanic Black and Hispanic patients at risk are less likely to have an Lp(a) order. Individuals residing in socioeconomically deprived neighborhoods and on Medicaid are also less like have Lp(a) order. Lp(a) orders depend on the type and number of patients’ diagnoses.

## Introduction

High levels of Lp(a) are causal for the development of atherosclerotic cardiovascular disease (ASCVD) and atherothrombosis as shown by epidemiological, Mendelian Randomization, and genome wide association studies (GWAS)^1^. Lipoprotein (a) [Lp(a)] levels are mostly regulated by genetics and it’s the most common genetic dyslipidemia. There are well-described mechanisms linking high levels of Lp(a) and atherosclerosis, including complement activation, inflammatory, and coagulation pathways^1–3^. Although most studies have been performed in Caucasian cohorts, there are well established reports on racial differences in Lp(a) levels. The UK biobank data showed significant variations in Lp(a) concentrations across racial subgroups, with highest levels in South Asians and Blacks^4^. Black patients were also shown to have the highest levels in the NOMASS study^5^. Lp(a) levels examined in the Atherosclerosis Risk in Communities (ARIC)^6^ and Multi-Ethnic Study of Atherosclerosis (MESA)^7^ studies, showed that in all racial/ethnic groups, including Hispanics, high Lp(a) levels were linked to an increase in ASCVD. Additionally, recent reviews have highlighted the role of structural racism on cardiovascular outcomes^8^.

International guidelines (Canadian^9^ and ECC^10^) advocate for all individuals to have Lp(a) measured once in a lifetime. Although some United States (ACC/AHA)^11^ guidelines suggest using Lp(a) levels as a ‘risk enhancer’, they do not recommend measuring Lp(a) levels in all. In the AHA scientific statement on Lp(a), the authors encourage identifying those with high Lp(a) early and suggested cut off values as ≥50 mg/dL or 125 nmol/L^11^. Particularly, this applies to those with premature atherosclerosis, persons at high risk for ASCVD and those with significant family history of ASCVD. The goal is to identify individuals and family members who would benefit from more intensive disease prevention, including LDL-C lowering^11^. The most recent update to the National Lipid Association (NLA) statement recommends, for the first time in the US, to measure Lp(a) in all adults once in a lifetime^12^. The aims of our study are to assess how racial/ethnic and socioeconomic factors impact Lp(a) ordering practices in a large urban academic institution. We hypothesized that patients’ race and ethnicity as well as their socioeconomic status—determined both by their environment (SES: socioeconomic score) and personal income-based dependency on Medicaid—impact the likelihood for Lp(a) orders.

## Methods

We used an institution-wide electronic medical record (EMR) database search of Columbia University Irving Medial Center (CUIMC) hospital and clinic system between February 2020 and July 2023 using Tripartite Request Assessment Committee (TRAC) warehouse. CUIMC has started using the Epic platform for EMR in 2020, so we limited our retrospective search to these dates to avoid combining data from different software platforms. This data request includes security safeguards and approval from the Institutional Review Board (IRB# AAAU3678).

We queried the system for unique adult patients who were recorded to have least one International Classifications of Disease (ICD-10) diagnosis of ASCVD or resistant hyperlipidemia (defined as LDL-C >160 mg/dL while on statin therapy). ICD-10 diagnoses included: familiar hyperlipidemia (FH, E78.01), atherosclerotic coronary artery disease (CAD, I25.10), myocardial infarction (MI, I21.9), peripheral arterial disease (PAD, I73.9), carotid artery disease(I65.2), cerebrovascular incident (I63, including occlusion and stenosis of cerebral and precerebral arteries, resulting in cerebral infarction), aortic valve stenosis (AS, I35.0), family history of elevated Lp(a) (Z83.430), and family history of ASCVD (Z82.49). This population was chosen based on the ACC/AHA ‘risk enhancer’ recommendations for identifying those at risk^11^. We assessed whether these individuals had a signed laboratory order for Lp(a) and whether this order was subsequently completed by the patient. We extracted additional data to help assess socioeconomic determinants, which included patient demographics, race/ethnicity, presence of income dependent health coverage (Medicaid), and primary residency zip codes to link to neighborhood measures of SES. Medicaid coverage was used as a marker of personal income, as this benefit is awarded based on low-income eligibility. There are no clear Lp(a) testing insurance guidelines and advanced lipoprotein testing is generally not covered by public insurance, so we did not include coverage as a variable. Medicaid eligibility was used only as a marker of patient’s low income. However, we provide patient insurance information in **Supplemental Table 6**. We also collected test ordering providers, their level of training and associated clinical departments.

### Neighborhood SES

The Yost Index is a composite score of 7 measures of neighborhood-level education, income, housing, and employment, derived from the United States Census Bureau. Measures are weighted and reported as a percentile score from 1 (most affluent) to 100 (most deprived) ^13^. The Yost Index assigns scores at the census tract level. We linked census tracts to ZIP codes using a previously validated census tract to ZIP code cross-walk^14^. Within our population, patients were categorized into tertiles based on their score. The first tertile (the group with the lowest Yost score, labelled as ‘lowest SES’) is the 33^rd^ percentile, and the third tertile (the group with the highest Yost scores, labelled as ‘highest SES’) corresponds to the 66th percentile. Consequently, patients with the lowest SES reside in the most socioeconomically advantaged community, whereas patients with the highest SES -- in the most deprived.

## Statical Analysis

Extracted TRAC data was cleaned from duplicate visits, to ensure that only one visit and Lp(a) order per patient were recorded. Patients with no specified gender or US zip code (N=26) were excluded from analysis, **Supplemental Figure 1**. Most recent available LDL-C and Lp(a) levels were extracted to calculate population characteristics. Normally distributed variables were summarized by means and standard deviations (SD) while Lp(a) levels were summarized by median and interquartile range. Chi-squared test was used to compared Lp(a) ordering frequencies between sexes. Kruskal-Wallis test and Mann-Whitney test was used to compare median Lp(a) levels among different races/ethnicities and sexes, respectively. Poisson regression model was used to obtain associations between variables of interest (diagnoses, race/ethnicity, SES, Medicaid eligibility) and outcomes (Lp(a) orders). P<0.05 was considered statistically significant. All data were analyzed using standard R software (Version 4.2.2).

## Results

### Cohort Characteristics

The general CUIMC adult patient population between February 2020 and July 2023 comprised of approximately 1,265,646 patients and only 0.32% had Lp(a) ordered with 74.8% completion rate. From this cohort, we collected data on 56,833 (4.5%) adult individuals who were diagnosed with either ASCVD or resistant hyperlipidemia (LDL>160mg/dL while on statin therapy).

Among those with elevated LDL-C levels on statin treatment, 6.4% were not diagnosed with ASCVD, while 28.1% had high Lp(a) levels. The rate of Lp(a) ordering was higher (4.0%) in our study cohort compared to the general patient population queried. The rate of test completion after the order was placed was similar in both study groups.

The baseline characteristics of the population are presented on **Table 1**. The most common ASCVD diagnosis was coronary artery disease (55.8%), followed by peripheral arterial disease (18%) and carotid stenosis (16.5%) with 21% patients having at least two, and 4.9% had at least three concurrent diagnoses. As expected, the majority (82.3%) of the cohort were taking statins. We found that 28.4% of our study population had high Lp(a) (≥125 nmol/L or 50 mg/dL). Non-Hispanic Black patients had the highest median Lp(a) levels, followed by Hispanic and non-Hispanic White patients as assessed in both mg/dL (p<0.001) or nmol/L units (p<0001), **Figure 1, and Supplemental Table 2**. While most patients with available Lp(a) levels were tested only once, 13.1% had two or more recorded Lp(a) values during the study period.

**Figure 1.**
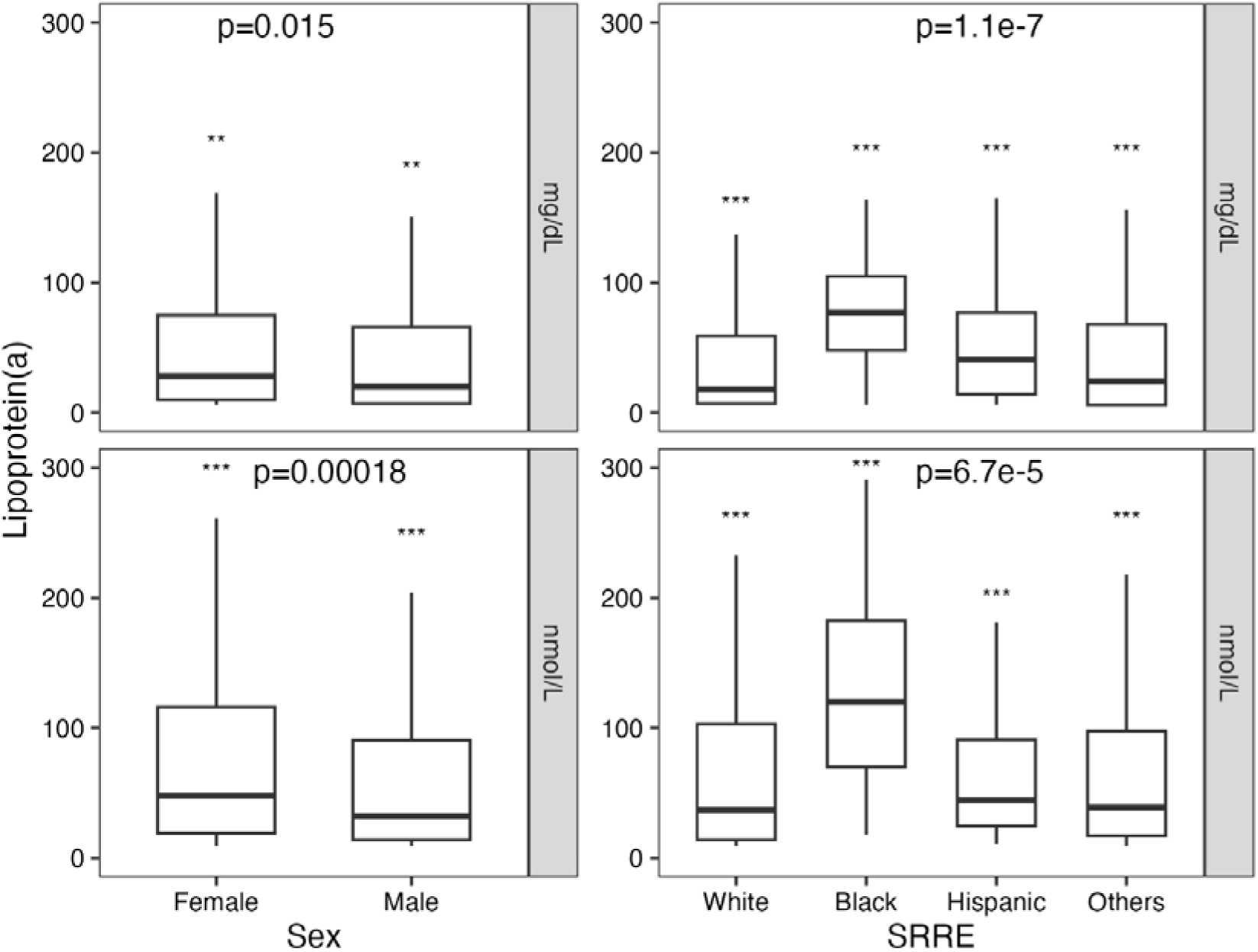
Lp(a) level differences by sex and race/ethnicity. p< 0.05 represents statistical significance; box plot represents comparison between median Lp(a) levels; Kruskal-Wallis and Mann-Whitney tests was used to compare Lp(a) level differences by sex and race/ethnicity, respectively.

**Table 1.**
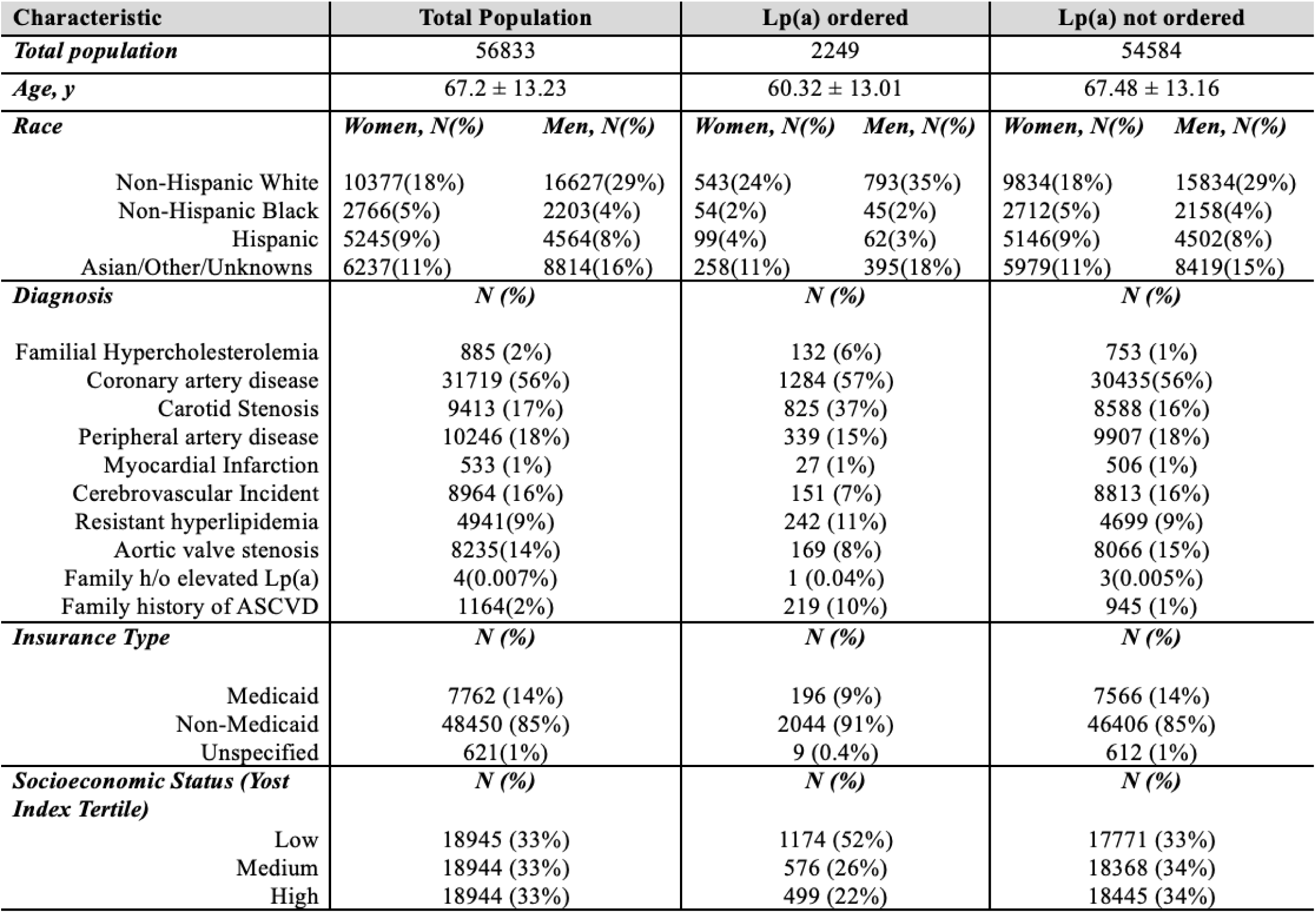
Population characteristics. ± represents Mean and Standard Deviation; y, years; resistant hyperlipidemia, LDL>160 mg/dL while taking statins; N=sample size; %, percentage from the total population; SES, socioeconomic score; Lp(a) *order* represents doctors’ prescription for Lp(a) measure; Lp(a) *tested* represents a completed test with a value

| Characteristic | Total Population |  | Lp(a) ordered |  | Lp(a) not ordered |  |
| --- | --- | --- | --- | --- | --- | --- |
| *Total population* | 56833 |  | 2249 |  | 54584 |  |
| *Age, y* | 67.2 ± 13.23 |  | 60.32 ± 13.01 |  | 67.48 ± 13.16 |  |
| *Race* | *Women, N(%)* | *Men, N(%)* | *Women, N(%)* | *Men, N(%)* | *Women, N(%)* | *Men, N(%)* |
| Non-Hispanic White | 10377(18%) | 16627(29%) | 543(24%) | 793(35%) | 9834(18%) | 15834(29%) |
| Non-Hispanic Black | 2766(5%) | 2203(4%) | 54(2%) | 45(2%) | 2712(5%) | 2158(4%) |
| Hispanic | 5245(9%) | 4564(8%) | 99(4%) | 62(3%) | 5146(9%) | 4502(8%) |
| Asian/Other/Unknowns | 6237(11%) | 8814(16%) | 258(11%) | 395(18%) | 5979(11%) | 8419(15%) |
| *Diagnosis* | *N (%)* |  | *N (%)* |  | *N (%)* |  |
| Familial Hypercholesterolemia | 885 (2%) |  | 132 (6%) |  | 753 (1%) |  |
| Coronary artery disease | 31719 (56%) |  | 1284 (57%) |  | 30435(56%) |  |
| Carotid Stenosis | 9413 (17%) |  | 825 (37%) |  | 8588 (16%) |  |
| Peripheral artery disease | 10246 (18%) |  | 339 (15%) |  | 9907 (18%) |  |
| Myocardial Infarction | 533 (1%) |  | 27 (1%) |  | 506 (1%) |  |
| Cerebrovascular Incident | 8964 (16%) |  | 151 (7%) |  | 8813 (16%) |  |
| Resistant hyperlipidemia | 4941(9%) |  | 242 (11%) |  | 4699 (9%) |  |
| Aortic valve stenosis | 8235(14%) |  | 169 (8%) |  | 8066 (15%) |  |
| Family h/o elevated Lp(a) | 4(0.007%) |  | 1 (0.04%) |  | 3(0.005%) |  |
| Family history of ASCVD | 1164(2%) |  | 219 (10%) |  | 945 (1%) |  |
| *Insurance Type* | *N (%)* |  | *N (%)* |  | *N (%)* |  |
| Medicaid | 7762 (14%) |  | 196 (9%) |  | 7566 (14%) |  |
| Non-Medicaid | 48450 (85%) |  | 2044 (91%) |  | 46406 (85%) |  |
| Unspecified | 621(1%) |  | 9 (0.4%) |  | 612 (1%) |  |
| *Socioeconomic Status (Yost Index Tertile)* | *N (%)* |  | *N (%)* |  | *N (%)* |  |
| Low | 18945 (33%) |  | 1174 (52%) |  | 17771 (33%) |  |
| Medium | 18944 (33%) |  | 576 (26%) |  | 18368 (34%) |  |
| High | 18944 (33%) |  | 499 (22%) |  | 18445 (34%) |  |

### Lp(a) Measurements across professional health specialties

Cardiologists comprised most of the ordering providers (86.8%), followed by Internal Medicine (9.52%), and Neurology (0.44%). Attending physicians were the main ordering professionals (88.8%), followed by mid-level providers, such as Nurse Practitioners and Physician Assistants (7.65%). Medical trainees (residents and students) ordered 2.36% of Lp(a) tests, **Supplemental Table 1A, B**.

### Relationship between ASCVD diagnoses and Lp(a) ordering

Individuals with diagnoses of familial hypercholesterolemia had the highest rate of Lp(a) orders (14.9%) follow by those with carotid stenosis (8.7%), myocardial infarction (5.1%), resistant hyperlipidemia (4.9%), coronary artery disease (4.0%), peripheral arterial disease (3.3%), aortic valve stenosis (2.1%), and cerebrovascular incident (1.7%). Ranking of diagnosis most likely to result in an Lp(a) order are illustrated in **Figure 2**, with carotid stenosis, family history of ASCVD and familial hypercholesterolemia more likely to have an Lp(a) order compared to others when considered together. Patients with greater number of ASCVD diagnoses (two, three, four or five) were significantly more likely to have an Lp(a) order compared to those with only one (p<0.001), **Supplemental Table 3**.

**Figure 2.**
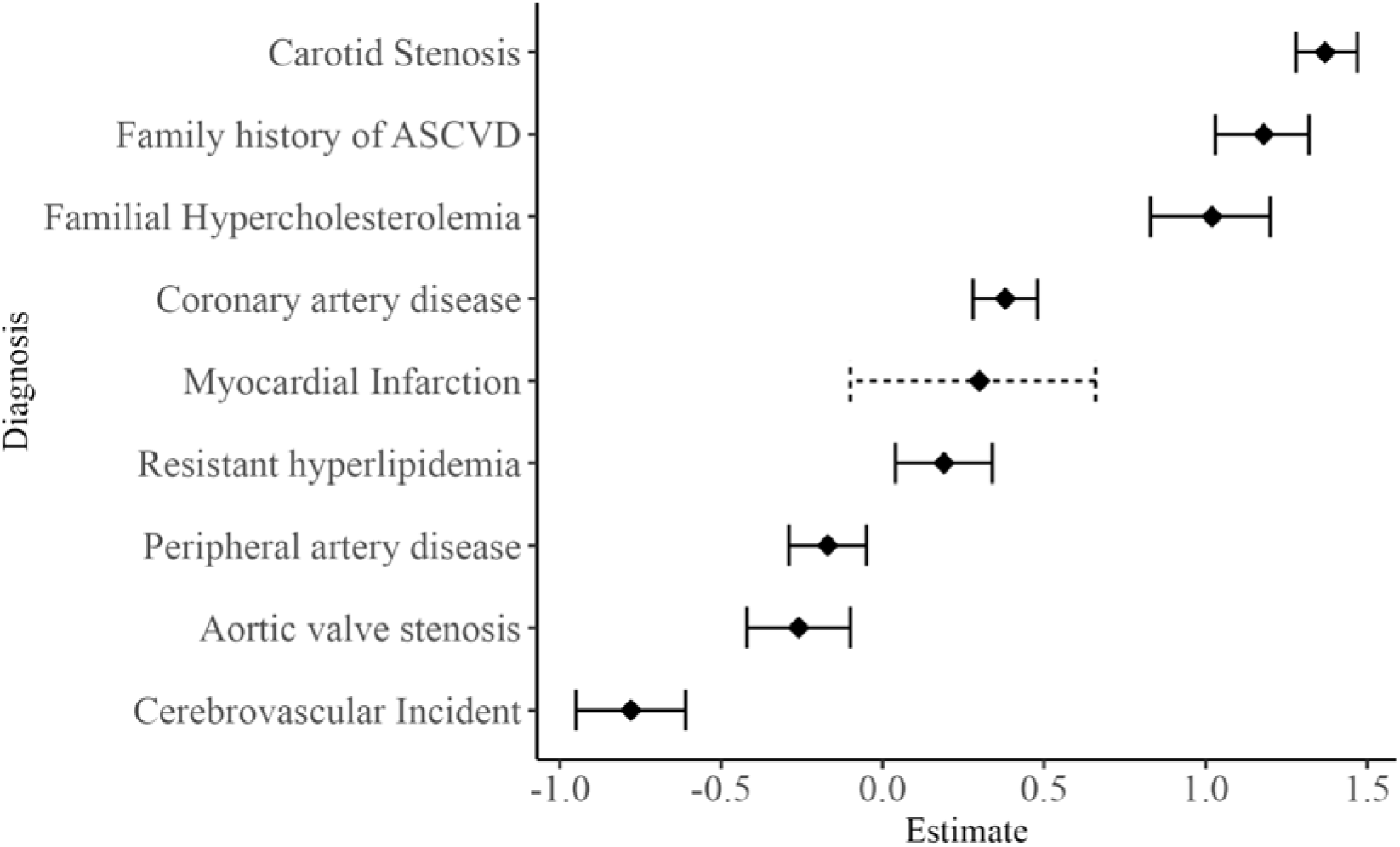
Lp(a) ordering pattern based on patient’s diagnosis profile. A Poisson regression model, adjusted for age and sex, was used to obtain beta estimates. Plot represents confidence interval of ranks.

### Lp(a) ordering practices in young patients (<40 years old) with ASCVD

Of all the patients with at least one ASCVD diagnosis, 3.32% (N=1,886) were under the age of 40 and 29.8% of this younger population had a history of a cardiovascular event (cerebral infarction or acute myocardial infarction). Additionally, 23% had LDL >160mg/dL while on statin therapy. Only 8.8% of patients with a premature cardiovascular event had an Lp(a) order and 73.9% of this group completed the test, **Supplemental Table 4**. *Younger patients* with diagnoses of familial hypercholesterolemia, coronary artery disease, and carotid disease were significantly more likely to have an Lp(a) order compared to the *older patients (*≥ *40 years old)* with the same diagnosis, **Supplemental Table 5**.

### Relationship between race/ethnicity, sex and Lp(a) ordering

Non-Hispanic Black (RR=0.32, 95% CI 0.26 - 0.40, p<0.001) and Hispanic patients (RR=0.28, 95% CI 0.23 - 0.32, p<0.001) were less likely to have an Lp(a) order compared to their non-Hispanic White counterparts. This association persisted, even when stratifying by neighborhood SES **(Table 2A).** Women had higher levels than men, **Figure 1**. Although there were no sex differences in Lp(a) ordering in the general population, when each race was assessed separately, Hispanic women were found to be more likely to have an Lp(a) order compared to Hispanic men (4% vs 3%, p=0.04).

**Table 2A.** Effects of race/ethnicity on Lp(a) ordering practices, stratified by socioeconomic status.

| SES | % with Lp(a) order | RR (95% CI) | P-value | Race/Ethnicity | RR (95% CI) | P-Value |
| --- | --- | --- | --- | --- | --- | --- |
| **Lowest SES** | 4.5% | 1.0 | - | *Non-Hispanic White* | 1.0 | - |
|  |  |  |  | *Hispanic* | 0.74 (0.53, 1.00) | 6.54e-02 |
|  |  |  |  | *Non-Hispanic Black* | 0.66 (0.43, 0.96) | 3.87e-02 |
|  |  |  |  | *Others* | 0.96 (0.84, 1.09) | 0.52 |
| **Middle SES** | 2.1% | 0.48 (0.43, 0.53) | 6.25e-47*** | *Non-Hispanic White* | 1.0 | - |
|  |  |  |  | *Hispanic* | 0.36 (0.24, 0.52) | 1.32e-07*** |
|  |  |  |  | *Non-Hispanic Black* | 0.35 (0.22, 0.53) | 5.23e-06*** |
|  |  |  |  | *Others* | 0.79 (0.65, 0.94) | 0.01 |
| **Highest SES** | 1.7% | 0.39 (0.35, 0.43) | 2.81e-70*** | *Non-Hispanic White* | 1.0 | - |
|  |  |  |  | *Hispanic* | 0.22 (0.17, 0.28) | 2.61e-33*** |
|  |  |  |  | *Non-Hispanic Black* | 0.28 (0.21, 0.38) | 1.30e-16*** |
|  |  |  |  | *Others* | 0.58 (0.47, 0.72) | 8.6e-7 |

### Relationship between SES, personal income and Lp(a) ordering

Patients residing in the more deprived neighborhoods were less likely to have an Lp(a) order compared to those residing in more affluent neighborhoods (using lowest SES as the reference group, middle SES: RR=0.48, 95% CI 0.43- 0.53, p<0.001; highest SES: RR=0.39, 95% CI 0.35 - 0.43, p<0.001). This was also true when stratified by race/ethnicity **(Table 2B)**. SES did not significantly impact whether the patients completed the test once the order was placed by the provider. Patients with Medicaid were significantly less likely to have an Lp(a) order compared to those without Medicaid (2.5% vs 4.2%, RR=0.40, 95% CI 0.34 - 0.46, p<0.001). This association also persisted after stratifying by race/ethnicity.

**Table 2B.** Effects of socioeconomic status on Lp(a) ordering practices, stratified by race/ethnicity. ***Statistical significance: p <0.001; SES, socioeconomic status; Poisson regression model adjusted for age and sex; RR = rate ratio (95% Confidence Interval), ‘non-Hispanic White’ and ‘highest SES’ serve as reference groups; ‘highest SES’ score corresponds to the most deprived neighborhood, ‘lowest SES’ score corresponds to the most affluent neighborhood

| Race/Ethnicity | % with Lp(a) order | RR (95% CI) | P-value | Race/Ethnicity | RR (95% CI) | P-Value |
| --- | --- | --- | --- | --- | --- | --- |
| **Non-Hispanic White** | 2.35% | 1.0 | - | *Lowest SES* | 1.0 | - |
|  |  |  |  | *Middle SES* | 0.57(0.50,0.64) | 8.60e-19*** |
|  |  |  |  | *Highest SES* | 0.78(0.67,0.90) | 1.19e-3 |
| **Non-Hispanic Black** | 0.17% | 0.32 (0.26, 0.40) | 4.65e-27*** | *Lowest SES* | 1.0 | - |
|  |  |  |  | *Middle SES* | 0.30(0.17,0.55) | 6.82e-5*** |
|  |  |  |  | *Highest SES* | 0.33(0.21, 0.55) | 5.85e-6*** |
| **Hispanic** | 0.28% | 0.28 (0.23, 0.32) | 5.57e-53*** | *Lowest SES* | 1.0 | - |
|  |  |  |  | *Middle SES* | 0.28(0.17,0.44) | 1.49e-7*** |
|  |  |  |  | *Highest SES* | 0.23(0.16,0.33) | 4.29e-15*** |
| **Others** | 1.15% | 0.78 (0.71, 0.86) | 3.66e-07*** | - | - | - |

## Discussion

High Lp(a) is an important causal risk factor for development ASCVD^15^ due to its atherogenic, prothrombotic and proinflammatory effects and disease risk increases incrementally with Lp(a) levels^4^. This risk persists despite lifestyle modifications and other risk factor management, such as aggressive LDL-C lowering^16–18^. Despite the available evidence and guidelines, Lp(a) screening remains persistently low ^19–21^. We observed that only 0.32% of patients in our general population had an Lp(a) order during the study period. Even in patients at risk (who already have at least one ASCVD diagnosis), only 4.0% had an Lp(a) order, with an upward trend from 2020 to 2023. A study done in a mostly Caucasian population observed an overall Lp(a) testing rate of 0.25% in 2015, with an upward trend over subsequent years, but only reaching 0.34% by 2018^22^. The University of California health system reports Lp(a) testing rates of 0.3%, with slightly higher 1.8-2.9% in those with ASCVD diagnosis^19^ As expected, in our study, a patient who had multiple ASCVD diagnoses was more likely to be tested for high Lp(a) compared to those with only one diagnosis. After the physician placed the order, patient compliance with Lp(a) testing was comparable to that of a routinely ordered lipid panel in adults, at 73.9% and 82.6%, respectively^23^. Cardiologists by far surpass other specialties in ordering Lp(a) in this high-risk population.

Although literature consistently shows the correlation between high Lp(a), MI and calcific aortic valve stenosis^10,24,25^, our data suggests that patients with these two diagnoses were not the most likely to be tested for high Lp(a). Individuals with diagnoses of familial hypercholesterolemia had the highest rate of Lp(a) orders follow by those with carotid stenosis, myocardial infarction, and resistant hyperlipidemia. The latter could be due to consistent efforts by independent patient advocate foundations and leading institutions, which in the last 5-10 years have targeted community engagement and provided resources to both patients and health providers^26,27^.

Patients with resistant hyperlipidemia would benefit from having their Lp(a) measured and treated, as there is a high probability that the observed increased cholesterol is carried by Lp(a) particles^28^ ^29^. Moreover, elevated Lp(a) levels alone have demonstrated an increase in cardiovascular risk, which is further compounded when LDL-C levels are also high. In fact, Lp(a) testing was found to lead to an increased initiation lipid lowering therapy compared with LDL-C testing alone in another study^30^, highlighting the importance of Lp(a) testing in those who already have other dyslipidemias. We also found that providers were more likely to order Lp(a) in patients with high LDL if they were under 40 years old, compared to those who were older.

The new 2024 update to the 2019 NLA statement^12^ for use of Lp(a) in clinical practice, for the first time in the US, recommends measuring Lp(a) in all. NLA recognizes individuals with Lp(a) levels ≥125 nmol/L (50 mg/dL) as high risk, which acknowledges that Lp(a) is associated with an increased incidence of ASCVD even in the absence of a family history of heart disease.

Previous recommendations for Lp(a) screening primarily stemmed from studies based mostly on data from White cohorts, like the Framingham Offspring Study. However, current update to the NLA statement includes multiethnic datasets, such as the UK Biobank. Considerable racial/ethnic variations in Lp(a) levels have been reported in large studies^6,7,31^. Similar to these reports, Lp(a) levels in our cohort were significantly different across races/ethnicities regardless which units were used (nmol/L vs mg/dL). Non-Hispanic Black patients had the highest median Lp(a) levels, followed by Hispanic, with non-Hispanic White patients having the lowest levels. Despite this, we saw that non-Hispanic Black and Hispanics patients were less likely to be ordered an Lp(a) test compared to their non-Hispanic White counterparts. Additionally, the Northern Manhattan Stroke Study (which also assesses patients from Northern Manhattan as in our study) reported that both Black and Hispanic participants were found to have a greater stroke incidence and a greater predominance for intracranial atherosclerotic stroke than White participants^32^.

Social determinants of health encompass an individual’s economic stability, neighborhood and built environment, education access, health care access and their social and community relationships^33^. Socioeconomic status has been associated with several health outcomes including cardiovascular disease, mortality from all causes, infant mortality, mental disorders, and some types of cancer^34^. In our population, non-Hispanic Black and Hispanic patients (who also represented the majority of our Medicaid patients and lower-income neighborhoods) had higher Lp(a) levels. However, they were tested less frequently with each of these three socioeconomic factors (SES, personal income, race/ethnicity) appearing to have independent, negative effects.

While one might hypothesize that observed differences in levels are the result of selection bias from differences in testing frequency, one cannot explain the differences in testing frequency. If one wishes to improve equity in cardiovascular outcomes, we need to provide equitable care, and this begins with implementing similar screening and diagnosis practices^26^. Evidence supports that socioeconomic status affects healthcare for aging adults, particularly as financial resources are proportional to health status^35^. Additionally, the Framingham coronary heart disease scoring suggests that risk is underestimated for persons with lower SES^36^. To our knowledge, there have been no previous reports on differences in Lp(a) ordering based on income. Yet, it is clear that some SES cardiovascular risk differences result not from intrinsic patient factors but from differences in healthcare treatment. Recent data supports the integration of ethnic and socioeconomic factors in healthcare^8^.

A common misconception among clinicians regarding Lp(a), which might partially drive the lack of screening, is the perceived lack of therapeutic options for high Lp(a). However, there are several non-targeted options for patients with high Lp(a). Patients can obtain significant benefit from more aggressive lifestyle modifications and the maintenance of optimal risk factors throughout life^37^. Many might also benefit from more aggressive lipid lowering with statins and PCSK9 inhibitors^38^. Available apoB100 lowering therapies lower Lp(a) modestly^1,39^, and apo(a) lowering therapies using small interfering RNA (siRNA) and antisense technology are now in phase 2 and phase 3 trials^1,40^. Not screening those at risk closes the door for an opportunity to employ already available and soon to be available therapies for those at risk. It is important to note, that Lp(a) testing is not yet standardized, with laboratory results reporting Lp(a) in either units of particle concentration (nmol/L) or mass concentration (mg/dL). Although there is no widely acceptable conversion factor (and many institutions, including NLA recommend against it), both available values are acceptable and are predictive of ASCVD risk. Education, electronic health record system implementation policies, and lowering the cost of testing with appropriate insurance coverage could serve as the next steps in bridging the gaps in all-inclusive medical preventive care.

This study is not without limitations. Electronic medical records are not always complete, which results in having to exclude some patients with missing data from the analysis. Our data query is also limited to a 3.5-year period; however, this time frame was chosen to ensure the most comprehensive dataset using only a single merged health record system (Epic). It is possible that subjects may have had testing before the study period began, which was not captured in our analysis. Similarly, testing outside of the CUIMC would also not be captured in our query.

Observational EMR data does not allow us to point out specific reasoning why a test was or was not ordered for each individual. As there are no clear coverage policies available for each insurance, we did not assess whether the type of insurance affected the physician’s decision to order the rest. Medicaid was only used as a marker of personal low income. Lastly, this study was conducted at one institution in an urban environment, limiting generalizability.

## Data Availability

All data produced in the present work are contained in the manuscript

